# Investigation of an estimate of daily resting heart rate using a consumer wearable device

**DOI:** 10.1101/19008771

**Authors:** Allison Russell, Conor Heneghan, Subbu Venkatraman

**Affiliations:** Fitbit Inc., 199 Fremont Street, San Francisco, CA 94105

## Abstract

Resting heart rate is accepted as a valid overall indicator of cardiovascular risk and is a low-cost easy-to-measure metric that can be collected in both home and clinical settings. Wearable devices such as smartwatches and trackers can measure heart rate continuously using optical technology. This study reports initially on the agreement between the resting heart rate reported by a wearable device (Fitbit Charge HR and Fitbit Blaze) and a chest-based consumer electrocardiogram (Polar H7). In 45 subjects, the reported resting heart rates agreed within 0.89 bpm over a resting period of 5 mins. The same device and algorithm was then used in a population of 168 subjects (45.9± 15.6 yrs) who were asked to collect their resting heart rate for four days under seven different conditions and times (at wake sitting/lying down, pre-breakfast, pre-lunch, pre-dinner, and before bed sitting/lying down). The reported values were compared with the Daily Resting Heart Rate (DRHR) reported by the wearable device. The measurement lying down before wake were closest to the DRHR (mean error of 0.0 bpm with a 25-75^th^ percentile spread of ±3.5bpm. There was a significant circadian variation in the reported resting heart rate across subjects, with the highest values reported in the middle of the day. Within individual subjects, the median standard deviation of heart rate at rest was 3-5bpm even measured under nominally similar conditions over four days.

## Introduction

Resting heart rate (RHR) is a useful overall metric for cardiovascular health. Menown et al, provide a useful summary of evidence in the area [Menown 2016], in which RHR is seen as independently associated with cardiovascular and all-cause mortality in patients with hypertension and in those with established cardiovascular disease. Resting heart rate appears therefore to have utility as a general marker of cardiovascular risk. Despite its apparent utility however, the guidelines for definition of resting heart rate are not well defined, nor have there been many systematic reports on the effect of measurement conditions on its value. Albanese et al raised this issue in a recent paper [ALB16], in which they discussed the effect of various measurement conditions and whether a single value could be considered representative of an underlying physiological marker. We shall call such a marker the “Daily Resting Heart Rate” (DRHR) which conceptually is a single value per day which is broadly representative of the person’s cardiac physiology on that day. In theory, a clinician should be able to get a reliable and repeatable estimate of this DRHR using established clinical protocols and equipment such as 12-lead ECGs, Holter monitors, or even pulse counting. The typical guidelines for measurement of a “resting heart rate” are that the reading should be taken in an office setting, with the person at rest, and without recent exertion. There is an implicit assumption that a RHR measured in this way should be representative of the underlying DRHR. However, heart rate by its nature is quite labile so not surprisingly as we shall illustrate in this paper, the measurement of a heart rate at rest in the same subject will vary quite significantly over the course of a day, or under different measurement conditions, as was also noted by Albanese et al. [ALB16]. For example, some influences on the measurement of RHR could include whether the patient is in a sitting or supine position, the time of day at which the measurement is taken.

This topic takes on renewed importance since the recent appearance of consumer friendly smartwatches and trackers has made measurement of heart rate on a continuous basis more feasible, and many of these devices report a “resting heart rate” which can be tracked by users on a daily basis over long periods of time. However, there have been few reports on the validity of these resting heart rate measurements and how they would correlate with existing clinical measurements. In this paper, we report on the agreement of the daily resting heart rate reported by a consumer tracker (Fitbit Charge HR) with baseline measurements recorded using values of resting heart rate taken at a variety of times and under different conditions.

The daily resting heart rate (DRHR) is calculated by Fitbit by combining multiple periods of “heart rate at rest” measured throughout the day, where the accelerometer on the device has determined that the person is at rest, and has not recently been moving (within a previous 5 minutes window for example). To be included in the calculation, the heart rate signal estimated must be with a high confidence level (which means that the person has minimal movement during the period, and the optical signal is of sufficient quality to be considered good to excellent). For users who wear the Fitbit device at night, their sleeping heart rate is also included, and the formulation of Brage which maps lying heart rate to resting heart rate is used [BRA05]. In order to have sufficient data, a person must have at least 30 minutes of still periods with valid data during the day for a DRHR to be reported. In this way a single number is calculated every day for a person wearing the device. This DRHR estimate is calculated once per day after the person has completed their main sleep (e.g., if a person falls asleep on Wednesday evening 9/18 at 11PM, and wakes up at 7AM on Thursday 9/19, the value of DRHR reported for that day 9/19 is based on the previous day’s data plus the night’s sleep.

This study consisted of two parts. Firstly we confirmed the accuracy of the agreement between the heart rate measured by the wearable device referenced against an ECG based heart rate derived from a chest strap, over a period of five minutes when stationary (in 45 subjects). This confirms the validity of heart rate measurements obtained when a person is still. Secondly, we then asked 169 volunteers to measure their heart rate at rest under a variety of different conditions to examine the impact of time of day and position on measurement repeatability, and to compare with the calculated DRHR reported by the Fitbit system.

## Methods

The study was approved by Solutions IRB as Protocol 2016/07/1. Participants were recruited from a pool of volunteers who had previously expressed interest in participating in research studies with Fitbit and were already users of a Fitbit device (either Fitbit Charge HR or Fitbit Blaze). In Phase 1 of the study, 45 subjects were recruited to assess the agreement level of the Fitbit device with a reference chest band electrocardiogram based measurement (Polar H7, Polar Oy, Finland). These chest-strap devices have already been shown to obtain heart rates within 0.2 bpm of a reference clinical ECG when a subject is at rest [ETI19]. The Fitbit devices have an on-board heart rate algorithm which outputs an estimated heart rate every 1 second based on analysis of the optical signal measured by the green photoplethysmogram. The Fitbit algorithm also marks the heart rate estimate with a “confidence level” – if significant motion is detected or if the optical signal has a low SNR, these estimates are marked as less reliable. As noted above, only measurements of high confidence are included in the DRHR estimate. These subjects were requested to sit comfortably, and to rest for ten minutes prior to the measurement. The Polar heart rate system also provides a heart rate estimate at a 1 second interval. A” heart rate at rest “was defined for both devices by taking the median heart rate estimate over the 5-minute period of measurement The average error between the Polar HR estimate and the Fitbit estimate was calculated over the 5 minute window. The purpose of Phase 1 was to confirm that the Fitbit device can reliably measure a heart rate at rest.

In Phase 2, 169 participants (85 F) with ages ranging from 18-75 yrs were recruited and consented electronically. They were instructed to measure their resting heart rate seven times per day under specific conditions: (1) supine immediately after wake, (2) sitting in bed after waking, (3) sitting before breakfast, (4) sitting before lunch, (5) sitting before dinner, (6) sitting in bed before bedtime, and (7) supine in bed before bedtime (refer to Table 1 for a summary view). Measurement was taken by recording for 4 minutes with the wrist worn device after a 5 min rest period. Excess movement or poor-quality signal could affect the quality of the estimate so the Fitbit device has an internal labeling system in which a confidence level is assigned to the heart rate estimate. A confidence level of 3 is highest (no movement and good optical signal). Confidence levels 1 and 2 occur when some movement is present or the optical signal is noisier. Subjects were asked to refrain from exercise, eating or drinking 2 hours prior to each measurement and to flag any measurements where they deviated from the protocol. Each subject was requested to repeat this protocol over 4 successive days. Subjects were caffeine and medication free, with no known cardiac health issues. In theory each participant could contribute 28 measurements (7 per day over 4 days); in practice, valid data was collected from 162 unique users with an average of 24 measurements each. This translates to an overall measurement capture efficiency of 82.1%. Users did receive daily email reminders to take measurements which helped to contribute to a satisfactory overall measurement rate.

**Table 1:** Instructions for measurement of resting heart rate given to participants.

| Measurement | When | Position | Extra instruction |
| --- | --- | --- | --- |
| M1 | Immediately upon wake | Lying in bed | Find a moment at least 2 hours after exercising, eating or drinking. Find a quiet place to sit comfortably and relax for 5 minutes. After settling your heart rate, hold still for an additional 4 minutes. Record the start and end time of the 4 minute window. |
| M2 | Immediately after #1 | Sitting in bed |  |
| M3 | Before breakfast | Sitting out of bed |  |
| M4 | Before lunch | Sitting out of bed |  |
| M5 | Before dinner | Sitting out of bed |  |
| M6 | Immediately before bed | Sitting in bed |  |
| M7 | Immediately after #6 | Lying in bed |  |

## Results

Figure 1 illustrates the results from Phase 1 of the study in which the heart rate from 45 subjects was taken with both Polar and Fitbit devices, and compared for 5-minute resting windows. The resting heart rate for the comparison windows can be taken as either the median HR reported during the window, or the minimum. We believe the median is a more robust measurement as it is less prone to outliers. The figure also shows the impact of the confidence level (quality of the heart rate estimate). Confidence levels 1 and 2 occur when some movement is present or the optical signal is noisier. Using the difference between median heart rates as the target, the overall average error over the 45 subjects was 0.89 bpm, and 93% of subjects had errors < 2bpm, with a standard error of 0.12 bpm. This is similar to the result reported by Hagayegh at el, [HAG19] who observed a difference of 0.09 bpm. Therefore we conclude from Phase 1 that the Fitbit device can provide a robust estimate of resting heart rate.

**Figure 1:**
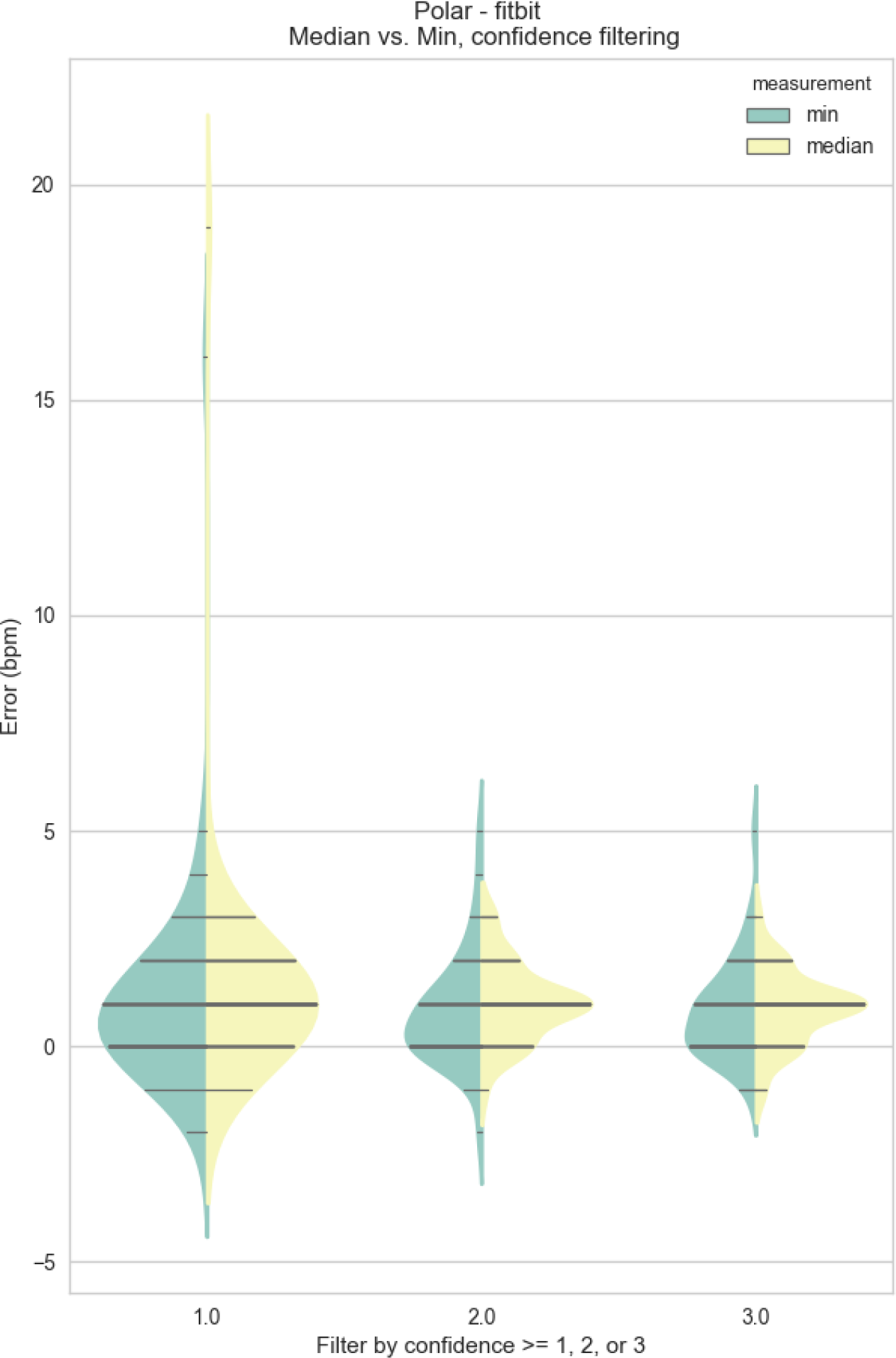
The distribution of the difference between the heart rate reported by the Polar device minus the heart rate reported by the Fitbit device. The error can be defined in two ways: a) taking the difference of the median heart rate reported by the devices over the 5 minute window, or b) taking the difference of the minimum values reported. This figure also shows the impact on error of only including the highest confidence measurements (least movement) which is when the Fitbit HR estimate confidence is 3. If the confidence level is only 1 or 2, then some movement has been detected and/or the optical signal quality if reduced. This would lead to a slightly higher error, particularly if a confidence level of 1 was used, so in Phase 2 of the study we restrict to higher confidence signals.

Phase 2 of the study was focused on the effect of real world measurement conditions on values of RHR, and the comparison of RHRs collected under those conditions with the underlying DRHR provided by the Fitbit device. Figure 2 shows the distribution of the participants in Phase 2 (age 45.9± 15.6 yrs). Recruitment was designed so that a broad range of users would be included.

**Figure 2:**
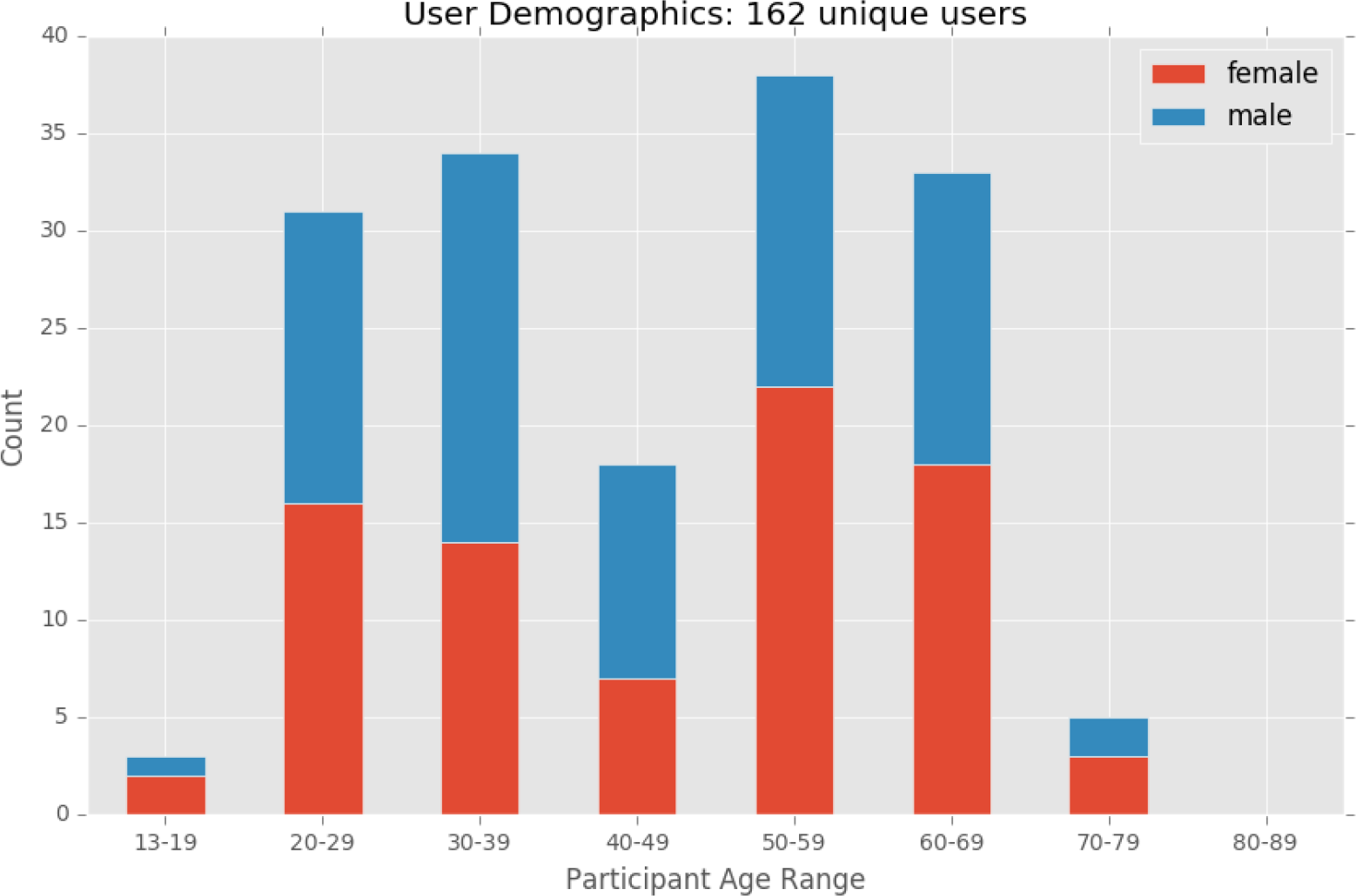
The distribution of participants in Phase 2 of the study (168 subjects).

Figure 3 shows the distribution of the differences between the “heart rate at rest” (HRaR) for that measurement window and the reported DRHR for that subject on that day. For example, if a user has a DRHR of 61 bpm for that day, and their reported “heart rates at rest for measurement windows [M1, M2, …, M7] were [59, 67, 65, 70, 65, 66, 61], then the differences for that user would be [-2, 6,4,9,4,4,0] bpm. The figure illustrates that there was significant diurnal variation in HRaR depending on the time of day of measurement. The lowest average HRaRs were recorded first thing in the morning in the supine position. Pre-lunchtime HRaR measurements were highest, and were 7 bpm higher on average than morning measurements. Within subjects, there was also considerable variability from day to day even under the same measurement conditions (the average intra-subject standard deviation for the morning measurement was 3.5bpm). The estimated resting heart rate (based on all-day measurements) reported by the wearable device was closest in value to the morning-supine measurement with a mean error of 0.0 bpm and a 25-75^th^ percentile spread of 3.5bpm. There are several interesting aspects to this figure: a) there is a well defined circadian rhythm with highest HRaRs reported during the active daytime period, and b) measurements taken lying down are consistently lower than when sitting as expected. Note that since this is a free living experiment, we do not have exact control of the circadian phase at which the HRaR is taken. Experimentally, it appears that the DRHR reported by the wearable device has the least amount of bias and measurement error when compared with ground truth measurements taken at M1 (first thing in the morning, lying down).

**Figure 3:**
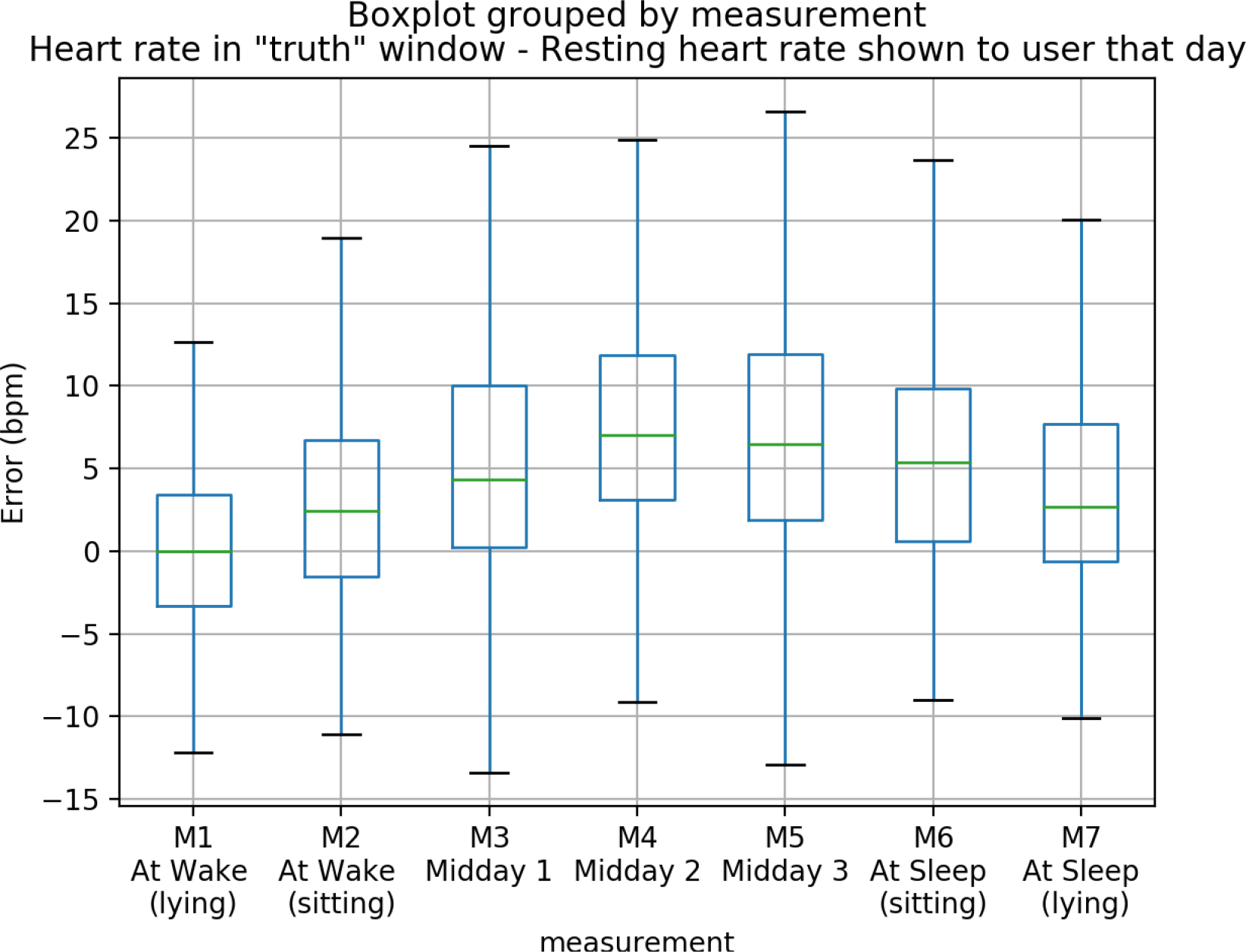
The influence of time of day and position (supine versus sitting) on the RHR measured. The differences between the “heart rate at rest” reported by the device for that measurement period minus the DRHR reported by the device (based on previous day’s measurements plus that night’s sleep). The box plot plus whiskers shows the median, 25-75th percentile and the 95th percentile values.

Fig. 4 gives an approximation of how labile the measurement of heart rate at rest is for individual users, even when taken under supposedly similar conditions. For example, if a single user took four measurements at the M3 measurement window which were [67, 72, 69, 75] then the standard deviation of their measurement at M3 is 3.5 bpm. Figure 4 shows the median standard deviation of the “heart rate at rest” taken at different time for users who took measurements at the designated time across all four days of the study. This illustrates again that the concept of single easily reproducible “resting heart rate” is not valid, but that in fact there is a reasonably large variation even under theoretically similar conditions. We believe that intuitively clinicians are aware that a single heart rate measurement at rest is not necessarily representative of an exact value of DRHR; however we are not aware of guidelines to clinicians on how many measurements they should average, or how spaced apart in time they should be.

**Fig. 4:**
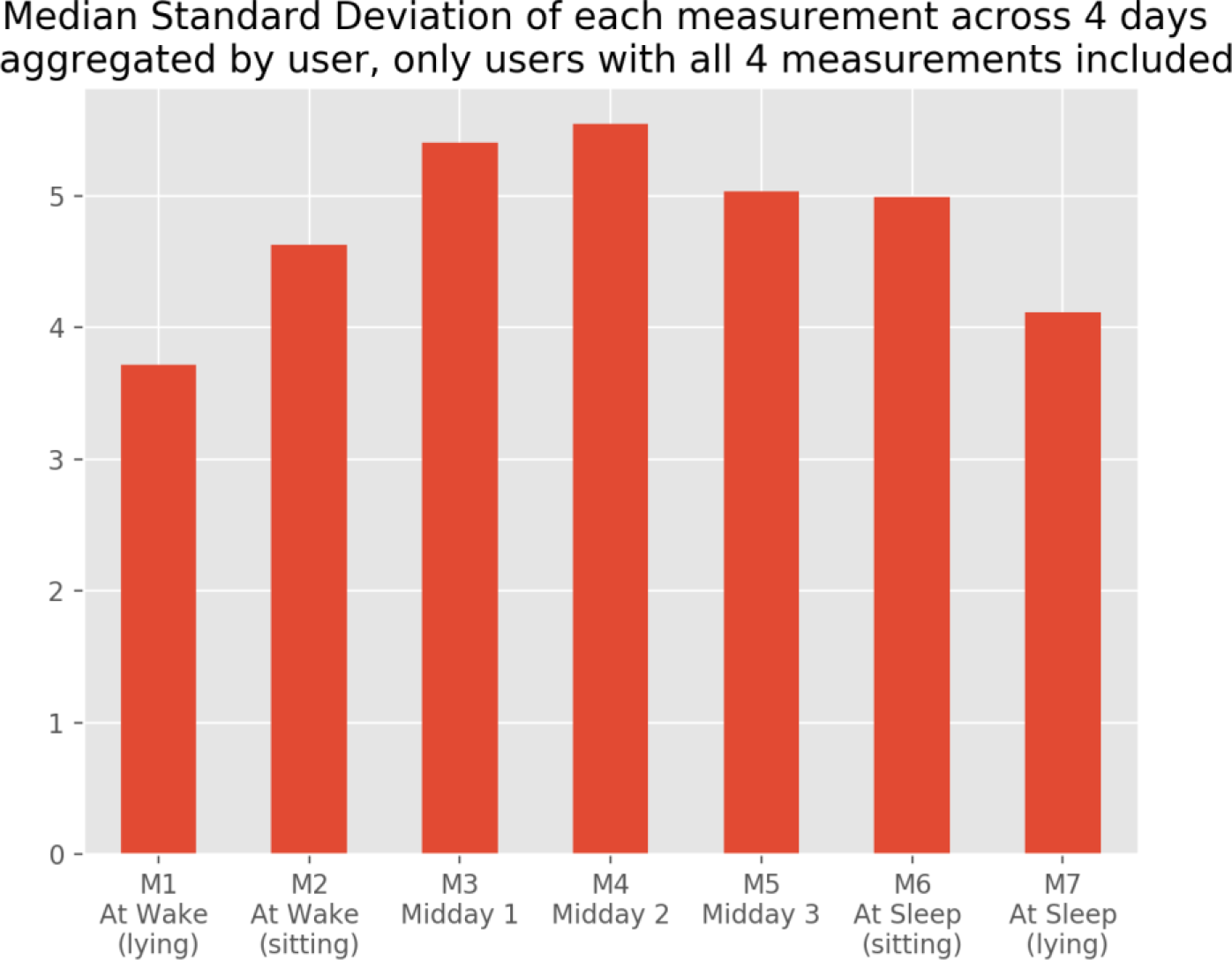
The median standard deviation of the “heart rate at rest” taken at different time for users who took measurements at the designated time across all four days of the subject.

## Discussion & Conclusions

The results on the accuracy of the heart rate measurement when at rest are consistent with previous results. In Haghayegh et al, they reported an average error of only 0.09bpm between the mean heart rate during sleep reported by the Fitbit Charge 2 device, and a 3-lead ECG reference [HAG19], which is similar to the observed value of 0.89 bpm over a 5-min observation window reported in this study. Resting heart rate measurements are highly dependent on time of day and measurement conditions, so caution should be taken clinically in relying on a single observation of RHR in an office visit. The observed circadian rhythm in resting heart rate is similar to that shown in Figure 2 of Wirz-Justice [WIR02], where they saw a change of about 4-5 BPM between the waking heart rate and the midday value. The daily resting heart rate reported by the wearable device used in this study was most similar to a heart rate obtained at waking up, while lying down. The DRHRs reported by the wearable device may have utility therefore in assessing the underlying resting heart rate of the wearer, and could also be used to potentially track longitudinal changes with a reasonable degree of validity.

## Data Availability

Interested researchers can contact the authors for data availability

